# Factors associated with cervical cancer stigma among married women living in urban Nepal

**DOI:** 10.64898/2026.10.28.26364684

**Authors:** Priyanka Timsina, Bandana Paneru, Manjari Shrestha, Asika Ghemosu, Bihungum Bista, Archana Shrestha

## Abstract

Cervical cancer stigma can affect perceptions of cancer, interactions with affected people, and engagement in preventive behavior. Evidence on factors associated with cervical cancer stigma among women in urban Nepal is limited. This study examined sociodemographic, reproductive and knowledge-related factors associated with cervical cancer stigma. We analysed baseline data from 310 women aged 30–60 years enrolled in a community-based, two-arm cluster-randomized controlled trial in Budanilkantha Municipality, Kathmandu, Nepal, between 15 June and 15 December 2022. Cervical cancer stigma was assessed using the validated Nepali version of the Cancer Stigma Scale. Multivariable generalized estimating equations (GEE) with a Gaussian distribution and identity link were used to examine associations, adjusting for age, ethnicity, religion, education, occupation, and annual household income. Results are reported as beta coefficients (β) and 95% confidence intervals (CIs). The mean age was 41.5 years, and 54.2% of participants were Janjati. Overall, 82.3% had heard of cervical cancer, 31.8% knew its signs and symptoms, and 1.6% had heard of Human Papilloma Virus. Most knew cervical cancer could be prevented (75.6%) and treated (90.1%). After adjustment, Janjati women (β = 0.311, 95% CI: 0.099, 0.523) and women from other ethnic groups, including Dalit and Madhesi (β = 0.411, 95% CI: 0.054, 0.768), had higher cervical cancer stigma scores than Brahmin/Chettri women. Formal education was associated with lower cervical cancer stigma scores (β = -0.359, 95% CI: - 0.554, -0.165). Homemakers (β = 0.263, 95% CI: 0.014, 0.513) and self-employed women (β = 0.309, 95% CI: 0.054, 0.564) had higher cervical cancer stigma scores than salaried women. Awareness of cervical cancer (β = -0.249, 95% CI: -0.437, -0.060), its signs and symptoms (β = - 0.245, 95% CI: -0.465, -0.025), preventability (β = -0.285, 95% CI: -0.554, -0.016) and treatability (β = -0.362, 95% CI: -0.635, -0.089) was associated with lower cervical cancer stigma scores. Age, religion, income, risk-factor awareness, menopause history, number of pregnancies and use of Intra Uterine Contraceptive Devices (IUCD) use were not significantly associated with cervical cancer stigma. Cervical cancer stigma among women in urban Nepal was associated with educational status, ethnicity, employment, and specific aspects of cervical cancer knowledge. Longitudinal and intervention studies are needed to determine whether improving cancer knowledge can effectively reduce cervical cancer stigma.

## Introduction

Cervical cancer remains a major global public health problem, particularly in low- and middle-income countries (LMICs). In 2022, around 604,000 new cases and 342,000 deaths from cervical cancer were reported globally. Nearly 90% of these burdens occur in LMICs where access to HPV vaccination, cervical cancer screening, early diagnosis, and treatment is limited [1]. Cervical cancer is largely preventable, and the World Health Organization (WHO) has set targets for HPV vaccination, screening, and treatment as part of its strategy to eliminate cervical cancer as a public health problem.

In Nepal, cervical cancer is the second most common cancer among women, with a crude incidence rate of 16.4 per 100,000 women and a mortality rate of 11.1 per 100,000 women [2]. Nepal has introduced several measures to improve prevention and early detection of cervical cancer through national guidelines and programs [3,4]. Despite these efforts, screening uptake remains very low, just 16% in community settings [5]. Lack of knowledge, embarrassment, limited access to services, concerns about privacy, and health-system barriers have been reported as important reasons for not using screening services [5,6].

Cancer-related stigma can have important consequences for cancer prevention, diagnosis, treatment and survivorship. Studies have shown that cancer-related stigma may be associated with delayed help-seeking, avoidance, social isolation, and reduced engagement with cancer-related services [7–9]. Stigma may also influence perceptions of cancer severity, personal responsibility and treatment, which are dimensions captured by the Cancer Stigma Scale (CASS) [10].

In Nepal, cancer-related stigma is likely shaped by the interplay of cultural norms, social attitudes, and religious beliefs. Previous studies have reported beliefs that cancer is inevitably fatal or contagious, as well as fears of social exclusion and misconceptions surrounding cancer [11]. Cervical cancer may carry additional stigma because of its association with HPV and sexual transmission. Misconceptions that HPV infection reflects multiple sexual partners or socially unacceptable sexual behavior may lead to embarrassment, fear of judgment, and reluctance to seek cervical cancer screening [6]. Mistrust in healthcare services may further compound these concerns and contribute to delayed or avoided care [12]. Importantly, evidence from Nepal suggests that stigma is associated with screening behavior: one study reported 23% lower cervical cancer screening coverage among women with higher levels of stigma compared with women with lower stigma [13].

Previous studies have shown that cancer stigma varies by age, gender, marital status, socioeconomic status, and family living arrangements [14,15]. Education and cancer-related knowledge may be particularly relevant to cancer stigma. Health literacy encompasses the ability to obtain, understand, appraise and use health information to make appropriate health decisions, and inadequate health literacy has been associated with poorer engagement with cancer screening and prevention [16,17]. Similarly, evidence from cancer communication research suggests that people with limited health literacy may experience greater difficulty navigating cancer prevention, screening, diagnosis and treatment [18].

Despite these findings, evidence on factors associated with cervical cancer stigma among women living in urban Nepal remains limited. Most existing Nepalese research has focused on overall cancer stigma in general [13,19]. These studies have mainly focused on sociodemographic and socioeconomic characteristics, while less attention has been given to reproductive factors and knowledge related to cervical cancer, its causes, prevention and screening. Given the role that reproductive experiences and knowledge may play in shaping perceptions of cervical cancer, examining these factors alongside sociodemographic characteristics may provide a more comprehensive understanding of stigma among women. Therefore, this study aimed to assess cancer stigma and examine its association with sociodemographic characteristics, reproductive characteristics, and cervical cancer-related knowledge among women aged 30–60 years living in an urban municipality of Nepal.

## Methods

### Study design and setting

We conducted a baseline analysis of a parent study that evaluated the effectiveness of a community-based intervention to reduce cancer stigma and improve cervical cancer screening-related outcomes [20]. The present analysis was restricted to data collected at baseline, before implementation of the intervention. The study was conducted in Budanilkantha Municipality, Kathmandu District, Bagmati Province, Nepal, between June 2022 and February 2023.

Budanilkantha municipality is divided into 13 administrative wards and has a diverse, predominantly urban population [21]. It is one of the largest municipalities in Nepal, with a total female population of 18,010 [21]. The major ethnic and social groups residing in the municipality include Janajati, Brahmin, Chhetri, Tamang, and Dalit communities [21].

In the parent study, wards served as clusters and were randomly allocated in a 1:1 ratio to the intervention or control group. Ward 3 was excluded before randomization because an existing cervical cancer screening program was operating in that ward. The remaining 12 wards were randomly allocated to six intervention and six control wards. From each ward, 25 women were taken. From each ward, 25 eligible women were recruited, resulting in a total sample of 310 women.

### Study Population

From 15 June 2022 and 15 December 2022, we enrolled 310 women in our study. Female community health volunteers (FCHVs) and elected female ward representatives were oriented to the study objectives, eligibility criteria, recruitment procedures, and their roles in community mobilisation. They disseminated information about the study and helped identify potentially eligible women. Eligibility was subsequently assessed by trained study staff. The inclusion criteria were (a) aged 30-60 years as recommended by National Guideline for Cervical Cancer Screening and Prevention in Nepal for cervical cancer screening 2010 [4] (b) married because the parent trial wanted to check effectiveness of stigma reduction intervention in cervical cancer screening uptake only married women would undergo screening (c) permanent residents of Budanilkantha Municipality. The exclusion criteria included (a) women with hearing impairment or mental health problems that prevent them from providing written informed consent, (b) pregnant or those less than six weeks postpartum at the time of data collection, (c) women who had resided in Budanilkantha for less than six months due to seasonal work (e.g., in brick kilns) or as visitors to the families of Budanilkantha residents, (d) women already diagnosed with cervical precancer, and (e) prior cancer affected women leading to hysterectomy.

### Data collection and measures

Data were collected through face-to-face interviews using a structured electronic questionnaire administered on tablets through Research Electronic Data Capture (REDCap) [22]. The questionnaire collected information on sociodemographic characteristics, reproductive and characteristics, knowledge regarding cervical cancer, cervical cancer screening practices and cervical cancer stigma.

Sociodemographic variables included age (years), ethnicity (Janjati, Brahmin/Chhetri, or other), educational attainment (no formal education or ≥1 year of formal education), occupation (homemaker, salaried employment, agriculture, daily wage labour, self-employment and others), and annual household income (US dollars). Reproductive variables included menopausal status (yes, no) and number of pregnancies (≤2 and ≥3). Knowledge-related variables assessed whether participants had previously heard of cervical cancer, as well as their awareness of its signs, symptoms and risk factors categorized as (yes/no/don’t know). Participants were also asked about their awareness of human papillomavirus (HPV) infection categorized as (yes/no/don’t know) and their knowledge of whether cervical cancer can be prevented and treated categorized as (yes/no/don’t know). In addition, participants were asked whether they had heard of cervical cancer screening categorized as (yes/no/don’t know). Due to the small number of responses in the “no” and “do not know” categories, these responses were combined into a single category (“no/do not know”) for analysis, with “yes” retained as the reference category.

We measured cervical cancer stigma using Cancer Stigma Scale (CASS)[10]; a validated tool in Nepali language [23]. CASS has 25 items assessing six domains: severity (4 items), personal responsibility (4 items), awkwardness (4 items), avoidance (4 items), policy opposition (4 items), and financial discrimination (5 items). Participants rated each item on a six-point Likert scale ranging from 1 (strongly disagree) to 6 (strongly agree). Severity domain assessed the perceived consequences of a cancer diagnosis and the likelihood of recovery, personal responsibility domain assessed how much a person’s actions are believed to contribute to their cancer, awkwardness domain assessed the comfort levels of cancer patients, avoidance domain assessed the extent to which people avoid cancer patients and maintain physical distance, the policy opposition domain assessed the perception of government and public responsibility for cancer patients’ treatment and care, financial discrimination domain assessed the extent to which cancer patients are expected to benefit from financial services such as banks and insurance. For each domain, item scores were summed and divided by the number of items in that domain to obtain a mean domain score. Thus, the possible score range was 1-6 for each domain. Five positively worded items in the awkwardness and policy opposition domains were reverse-coded before calculating the domain scores, such that higher scores consistently indicated greater cancer stigma [10]. An overall stigma score was calculated by taking the mean of all 25 items after reverse coding the positively worded items. The overall score therefore also ranged from 1 to 6, with higher scores indicating higher levels of perceived cancer stigma [10].

### Statistical Analysis

We performed all statistical analyses using Stata version 14.0 (StataCorp, College Station, TX, USA). We summarized categorical variables using frequencies and proportions, normally distributed numerical variables using means and standard deviations (SD), and non-normally distributed numerical variables using medians and interquartile ranges (IQR). We collected income data in Nepalese Rupees (NPR) and converted them to US Dollars (USD) using an exchange rate of 1 USD = 153.51 NPR, based on the average exchange rate on September 29, 2026. We calculated the cervical cancer stigma score as the mean of all 25 items [10].

As participants were clustered within wards, we assumed that women living in the same ward may have correlated outcomes due to shared sociodemographic, environmental, and cultural characteristics. To account for this clustering, generalized estimating equations (GEE) with a Gaussian distribution and identity link function were used to examine the associations between cervical cancer stigma scores and sociodemographic, reproductive, and cervical cancer knowledge-related characteristics. An exchangeable working correlation structure was specified, and robust variance estimators were used to obtain standard errors that account for within-ward correlation and remain valid even when the working correlation structure is mis specified.

Bivariable GEE models were first used to assess the unadjusted association between each exploratory variable, including sociodemographic, reproductive, and cervical cancer knowledge-related factors, and cervical cancer stigma score. A multivariable GEE model was then fitted to account for potential confounding by age, ethnicity, religion, educational status, occupation, and annual household income. The results are presented as regression coefficients (β) with 95% confidence intervals (CIs) and corresponding P-values.

### Ethics Statement

The Institutional Review Committee (IRC) of Kathmandu University School of Medical Sciences approved the study (IRC reference number: 42/22). We obtained written informed consent from participants who could read and write. For participants who were illiterate, we obtained verbal informed consent, and a witness chosen by the participant signed the consent form. We maintained confidentiality and ensured voluntary participation throughout the study. Participants could withdraw from the study at any time without any consequences.

## Results

Table 1 shows the sociodemographic, reproductive and knowledge related characteristics of 310 participants were included in the analysis. The mean age of the participants was 41.5 years. More than half of the participants belonged to the Janjati ethnic group (54.2%), followed by Brahmin/Chettri (41.6%) Most participants were Hindu (80.7%). Around 71.9% of the participants had received formal education. Nearly all participants were currently married (299, 96.5%). Homemakers constituted the largest occupational group (62.9%), followed by self-employed participants (18.1%) and those in salaried employment (10.0%). The median annual household income was USD 2,435 (IQR 1,563-3,908).

**Table 1.** Sociodemographic, reproductive and knowledge related characteristics of study participants.

| **Sociodemographic characteristics** | **Total (n=310)** |
| --- | --- |
| **Age, years, mean (SD)** | 41.52 (7.82) |
| **Ethnicity, n (%)** |  |
| Janjati | 168 (54.2) |
| Brahmin/Chettri | 129 (41.6) |
| Other | 13 (4.2) |
| **Religion, n (%)** |  |
| Hindu | 250 (80.65) |
| Non-Hindus | 60 (19.35) |
| **Educational status, n (%)** |  |
| No formal education | 87 (28.06) |
| Formal education | 223 (71.94) |
| **Occupation, n (%)** |  |
| Homemaker | 195 (62.90) |
| Salaried employment | 31 (10) |
| Daily wage labor | 10 (3.23) |
| Self-employed | 56 (18.06) |
| Other | 18 (5.81) |
| **Annual family income**<br>**median (IQR), USD** | USD 2,435 (IQR 1,563-3,908) |
| **Reproductive Characteristics** |  |
| **Menopause, n (%) (n=310)** | 58 (18.71) |
| **Number of pregnancies** |  |
| ≤ 2 | 165 (53.23) |
| ≥ 3 | 145 (46.77) |
| **Use of IUCD(n=252)** | 17 (6.69) |
| **Knowledge on cervical cancer** |  |
| Heard about cervical cancer(n=310) | 255 (82.26) |
| Aware about signs and symptoms(n=255) | 80 (31.37) |
| Aware about the risk factors(n=255) | 80 (31.75) |
| Heard about HPV infection(n=255) | 5 (1.96) |
| Can cervical cancer be prevented(n=255) | 192 (75.29) |
| Can cervical cancer be treated(n=255) | 228 (89.41) |
| Heard about cervical cancer screening(n=310) | 179 (57.74) |
| **Cervical cancer stigma score** | 3.10 (0.04) |

*SD, standard deviation; IQR, interquartile range; USD, United States dollar; IUCD,* *intrauterine contraceptive device; HPV, human papillomavirus.*

Regarding reproductive characteristics, 18.71% women reported having reached menopause. More than half of the participants, 53.23%, had experienced ≤2 pregnancies, while 46.77% had experienced ≥3 pregnancies. Among the 252 women who had not reached menopause, 6.69% reported having used an IUCD.

Most participants (82.26%) had heard of cervical cancer. However, only (31.37%) participants reported awareness of its signs and symptoms and its risk factors. Only (1.96%) participants had heard of HPV infection. Most participants knew that cervical cancer can be prevented (75.29%) and treated (89.41%). Awareness of cervical cancer screening was reported by (57.74%) participants. The mean cervical cancer stigma score was 3.10 (SD 0.04).

Table 2 presents the results of a multivariable generalized estimating equation (GEE) model examining the association between cervical cancer stigma and various sociodemographic factors. In the adjusted model ethnicity, educational status and occupation were statistically significant with cervical cancer stigma. Compared with participants from the Brahmin/Chettri women, Janjati women (β = 0.311, 95% CI: 0.099, 0.523) and women from other ethnic groups, including Dalit and Madhesi (β = 0.411, 95% CI: 0.054, 0.768) had higher cervical cancer stigma scores. Participants with formal education had lower cervical cancer stigma than those with no formal education (adjusted β = -0.359, 95% CI: -0.554, -0.165). Compared with participants in salaried employment, homemakers (β = 0.263, 95% CI: 0.014, 0.513) and self-employed women (β = 0.309, 95% CI: 0.054, 0.564) had higher cervical cancer stigma scores. Age (adjusted β = 0.009, 95% CI: -0.001, 0.018), religion (adjusted β = 0.113, 95% CI: -0.119, 0.346) and annual household income (adjusted β = -0.305, 95% CI: -0.635 to 0.032) were not statistically significantly associated with cervical cancer stigma in the adjusted model.

**Table 2.**
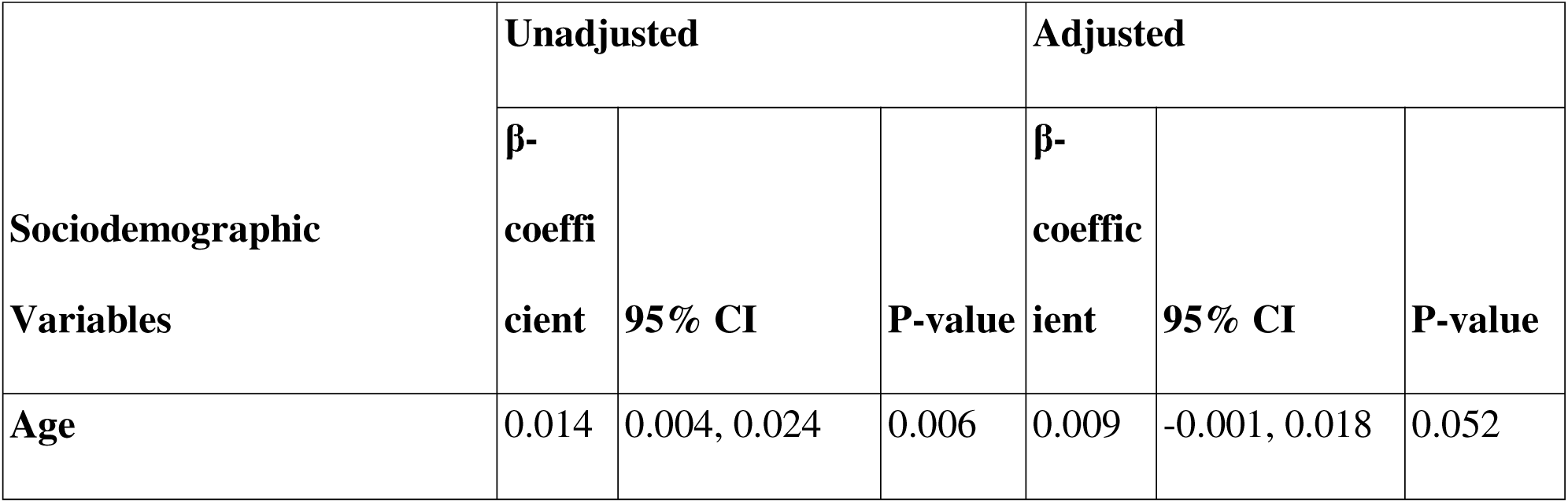
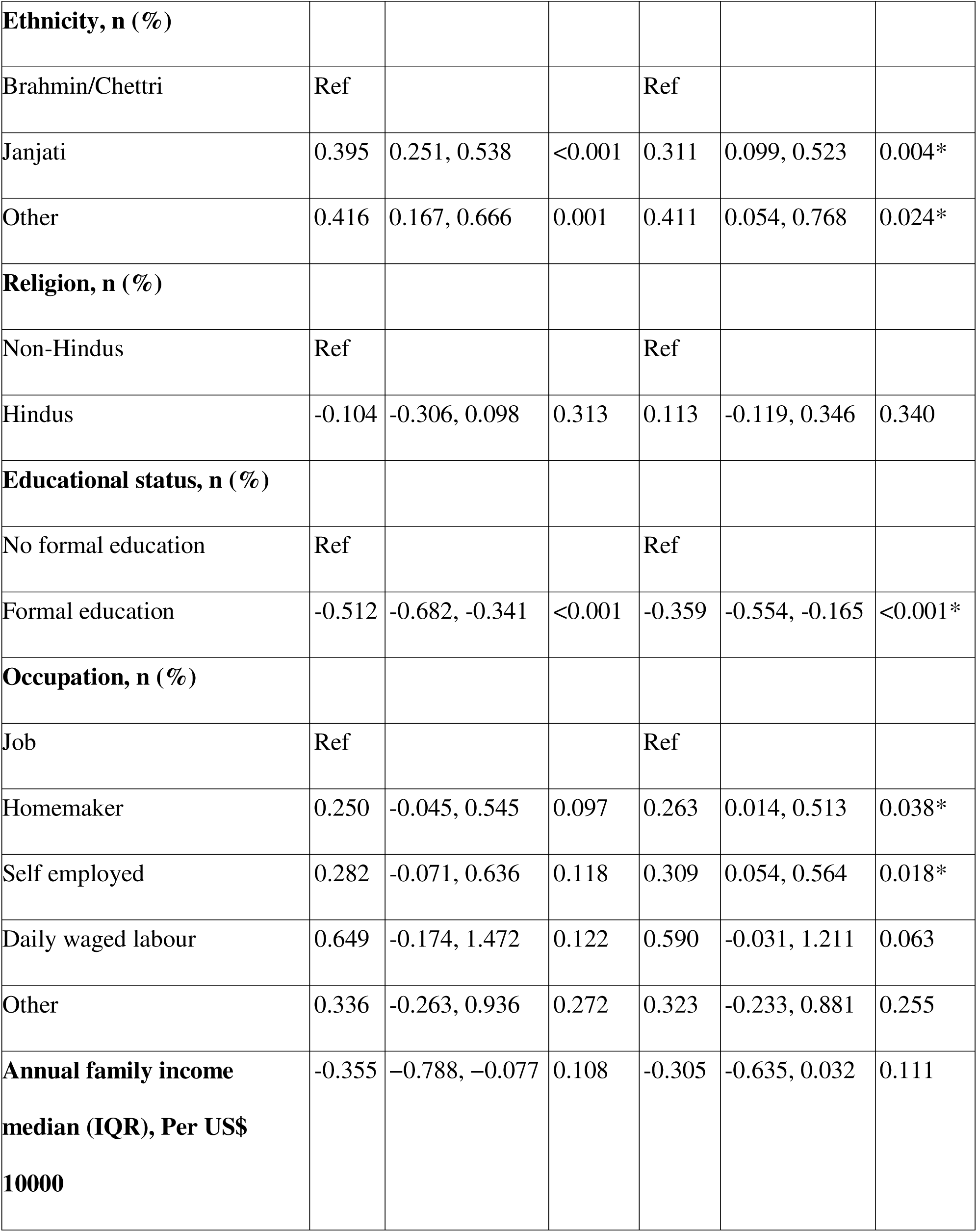
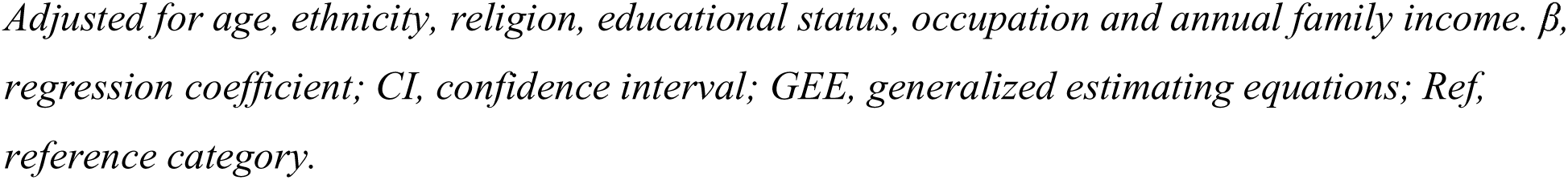
Association of sociodemographic variables with cervical cancer stigma score.

| Sociodemographic<br>Variables | Unadjusted |  |  | Adjusted |  |  |
| --- | --- | --- | --- | --- | --- | --- |
| | β-<br>coeffi<br>cient | 95% CI | P-value | β-<br>coeffic<br>ient | 95% CI | P-value |
| Age | 0.014 | 0.004, 0.024 | 0.006 | 0.009 | -0.001, 0.018 | 0.052 |
| **Ethnicity, n (%)** |  |  |  |  |  |  |
| Brahmin/Chettri | Ref |  |  | Ref |  |  |
| Janjati | 0.395 | 0.251, 0.538 | <0.001 | 0.311 | 0.099, 0.523 | 0.004* |
| Other | 0.416 | 0.167, 0.666 | 0.001 | 0.411 | 0.054, 0.768 | 0.024* |
| **Religion, n (%)** |  |  |  |  |  |  |
| Non-Hindus | Ref |  |  | Ref |  |  |
| Hindus | -0.104 | -0.306, 0.098 | 0.313 | 0.113 | -0.119, 0.346 | 0.340 |
| **Educational status, n (%)** |  |  |  |  |  |  |
| No formal education | Ref |  |  | Ref |  |  |
| Formal education | -0.512 | -0.682, -0.341 | <0.001 | -0.359 | -0.554, -0.165 | <0.001* |
| **Occupation, n (%)** |  |  |  |  |  |  |
| Job | Ref |  |  | Ref |  |  |
| Homemaker | 0.250 | -0.045, 0.545 | 0.097 | 0.263 | 0.014, 0.513 | 0.038* |
| Self employed | 0.282 | -0.071, 0.636 | 0.118 | 0.309 | 0.054, 0.564 | 0.018* |
| Daily waged labour | 0.649 | -0.174, 1.472 | 0.122 | 0.590 | -0.031, 1.211 | 0.063 |
| Other | 0.336 | -0.263, 0.936 | 0.272 | 0.323 | -0.233, 0.881 | 0.255 |
| **Annual family income**<br>**median (IQR), Per US\$**<br>**10000** | -0.355 | -0.788, -0.077 | 0.108 | -0.305 | -0.635, 0.032 | 0.111 |

Adjusted for age, ethnicity, religion, educational status, occupation and annual family income. $\beta$ , regression coefficient; CI, confidence interval; GEE, generalized estimating equations; Ref, reference category.

Table 3 presents the results of a multivariable generalized estimating equation (GEE) model examining the association between cervical cancer stigma and various reproductive factors. Menopause status(adjusted β = 0.751, 95% CI: 0.581, 2.084), number of pregnancies(adjusted β = 0.170, 95% CI: -0.005, 0.345) and IUCD use (adjusted β = -0.293, 95% CI: -0.693, 0.105) were not statistically significantly associated with cervical cancer stigma after adjustment for age, ethnicity, religion, educational status, occupation and annual family income.

**Table 3.** Association of reproductive variables with cervical cancer stigma score.

| Reproductive Variables | Unadjusted |  |  | Adjusted |  |  |
| --- | --- | --- | --- | --- | --- | --- |
| | β-<br>coefficient | 95% CI | P-value | β-<br>coefficient | 95% CI | P-value |
| **Number of pregnancies** |  |  |  |  |  |  |
| $\leq 2$ | Ref | | | Ref | | |
| $>3$ | 0.288 | 0.105, 0.471 | 0.002 | 0.170 | -0.005, 0.345 | 0.057 |
| **Menopause** |  |  |  |  |  |  |
| No | Ref |  |  | Ref |  |  |
| Yes | 0.266 | 0.094, 0.439 | 0.002 | 0.751 | -0.581, 2.084 | 0.269 |
| **Use of IUCD** |  |  |  |  |  |  |
| No | Ref |  |  | Ref |  |  |
| Yes | -0.258 | -0.679, 0.161 | 0.228 | -0.293 | -0.693, 0.105 | 0.149 |

*Adjusted for age, ethnicity, religion, educational status, occupation and annual family income. $\beta$ , regression coefficient; CI, confidence interval; GEE, generalized estimating equations; Ref, reference category. IUCD, intrauterine contraceptive device.*

Table 4 presents the results of a multivariable generalized estimating equation (GEE) model examining the association between cervical cancer stigma and knowledge related factors. After adjustment for age, ethnicity, religion, educational status, occupation, and annual household income, women who had heard about cervical cancer had lower stigma scores than those who had not heard about cervical cancer (adjusted β = -0.249, 95% CI: -0.437 to -0.061). Women who were aware of cervical cancer signs and symptoms had lower stigma scores than those who were not aware (adjusted β = -0.245, 95% CI: -0.465 to -0.025). Similarly, women who knew that cervical cancer could be prevented (adjusted β = -0.285, 95% CI: -0.554 to -0.016) or treated (adjusted β = -0.362, 95% CI: -0.635 to -0.089) had lower stigma scores than those who did not know. Women who had heard about cervical cancer screening had lower stigma scores than those who had not heard about screening (adjusted β = -0.207, 95% CI: -0.363 to 0.051). Awareness of cervical cancer risk factors was not significantly associated with stigma after adjustment (adjusted β = -0.183, 95% CI: -0.398 to 0.032).

**Table 4.**
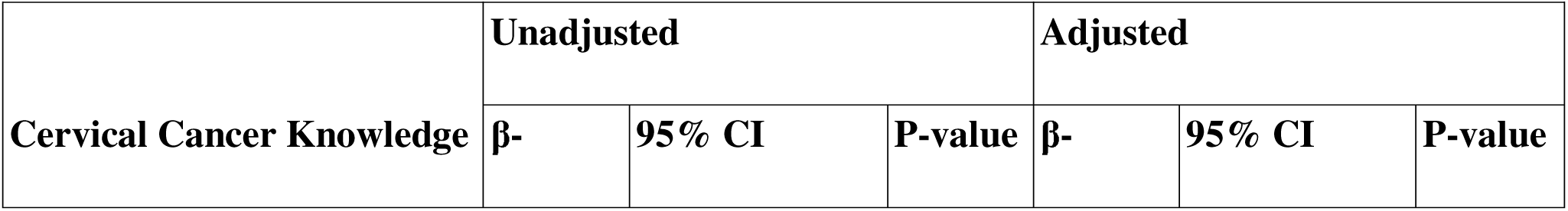
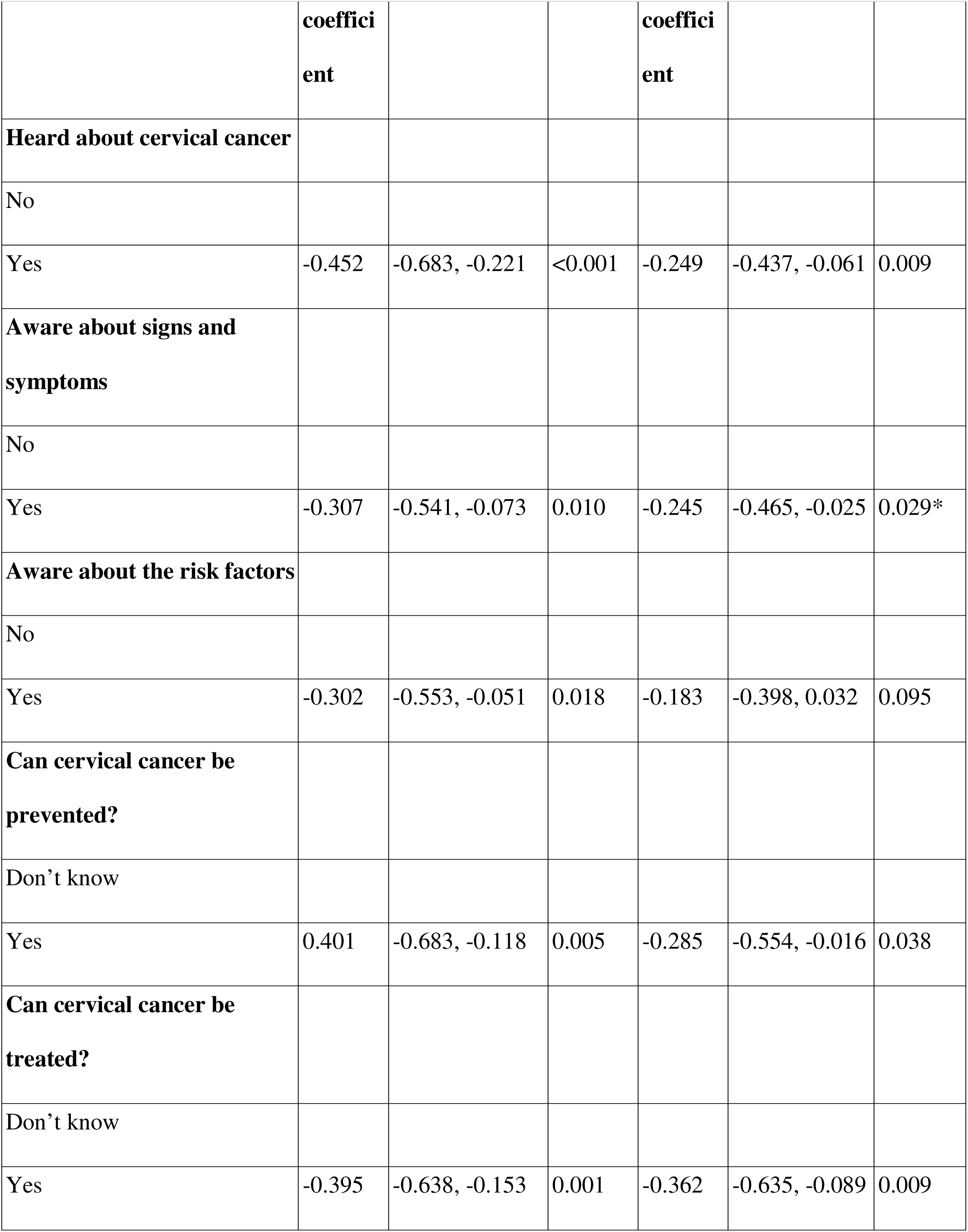
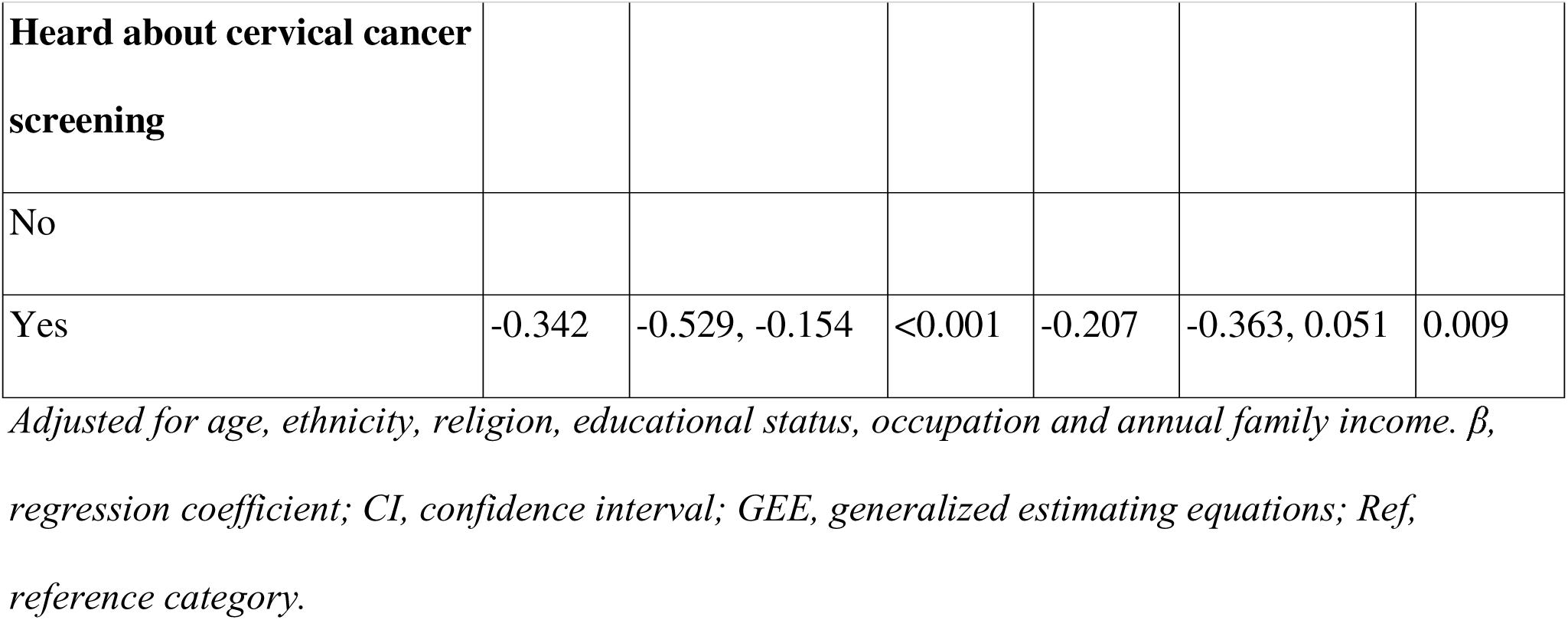
Association of cervical cancer knowledge with cervical cancer stigma score.

| Cervical Cancer Knowledge | Unadjusted |  |  | Adjusted |  |  |
| --- | --- | --- | --- | --- | --- | --- |
| | β- | 95% CI | P-value | β- | 95% CI | P-value |

|  | **coefficient** |  |  | **coefficient** |  |  |
| --- | --- | --- | --- | --- | --- | --- |
| **Heard about cervical cancer** |  |  |  |  |  |  |
| No |  |  |  |  |  |  |
| Yes | -0.452 | -0.683, -0.221 | <0.001 | -0.249 | -0.437, -0.061 | 0.009 |
| **Aware about signs and symptoms** |  |  |  |  |  |  |
| No |  |  |  |  |  |  |
| Yes | -0.307 | -0.541, -0.073 | 0.010 | -0.245 | -0.465, -0.025 | 0.029* |
| **Aware about the risk factors** |  |  |  |  |  |  |
| No |  |  |  |  |  |  |
| Yes | -0.302 | -0.553, -0.051 | 0.018 | -0.183 | -0.398, 0.032 | 0.095 |
| **Can cervical cancer be prevented?** |  |  |  |  |  |  |
| Don't know |  |  |  |  |  |  |
| Yes | 0.401 | -0.683, -0.118 | 0.005 | -0.285 | -0.554, -0.016 | 0.038 |
| **Can cervical cancer be treated?** |  |  |  |  |  |  |
| Don't know |  |  |  |  |  |  |
| Yes | -0.395 | -0.638, -0.153 | 0.001 | -0.362 | -0.635, -0.089 | 0.009 |
| **Heard about cervical cancer screening** |  |  |  |  |  |  |
| No |  |  |  |  |  |  |
| Yes | -0.342 | -0.529, -0.154 | <0.001 | -0.207 | -0.363, 0.051 | 0.009 |

*Adjusted for age, ethnicity, religion, educational status, occupation and annual family income. $\beta$ , regression coefficient; CI, confidence interval; GEE, generalized estimating equations; Ref, reference category.*

## Discussion

This study examined cervical cancer stigma and its sociodemographic, reproductive, and cervical cancer-related knowledge correlates among women aged 30–60 years living in an urban municipality of Nepal. Overall, stigma differed according to educational status, ethnicity, occupation, and several aspects of cervical cancer-related knowledge. Women with formal education reported lower stigma scores than those with no formal education, whereas women from Janjati and other ethnic groups including Dalit, Madhesi and others reported higher stigma scores than those from the Brahmin/Chettri group. Homemakers and self-employed women also reported higher stigma than women in salaried employment. In addition, women who had heard of cervical cancer and those who were aware of its signs and symptoms, prevention, treatment, and screening reported lower stigma scores. In contrast, awareness of cervical cancer risk factors was not significantly associated with stigma after adjustment. Taken together, these findings indicate that cervical cancer stigma may be shaped by both social characteristics and specific forms of disease-related knowledge, highlighting the importance of considering these factors when addressing stigma among women in urban Nepal.

The association between formal education and lower stigma is consistent with previous evidence from Nepal. Shrestha et al. found that women with no formal education reported higher stigma related to avoiding people with cancer and personal responsibility [1]. Similarly, Paneru et al. found that lower educational attainment was associated with higher levels of cancer stigma among women in semi-urban Nepal [2]. Studies from Pokhara and central Nepal have also reported limited knowledge of cervical cancer and screening, with higher education associated with better knowledge [3,4]. These findings suggest that differences in access to and understanding of health information may partly explain the lower stigma observed among more educated women in our study.

Ethnicity was independently associated with stigma. Janjati and other ethnic groups including Madhesi, Dalit and others had higher stigma scores than Brahmin/Chettri women. Previous research has reported differences in cancer stigma across ethnic groups [9,10]. Evidence from the original CASS development study found higher scores in several stigma domains among participants from ethnic minority groups in the United Kingdom [10]. Similarly, the English population-based study reported higher cancer stigma among ethnic minority participants[9]. However, findings from Nepal have been inconsistent. Paneru et al. did not find ethnicity to be independently associated with overall cancer stigma in their semi-urban Nepalese study [19]. In contrast, the Nepali CASS validation study identified differences in ethnicity across some stigma domains [23]. In our urban study, ethnicity was associated with cancer stigma, unlike in the previous semi-urban Nepalese study. This difference may be related to variations in the social and demographic characteristics of urban and semi-urban populations, as well as differences in access to health information and health services.

Occupation was also associated with stigma. Homemakers and self-employed women reported higher stigma than women in salaried employment. Evidence on the relationship between occupation and cancer stigma is limited and inconsistent. Previous studies in Nepal did not find a consistent independent association between occupation and overall cancer stigma [19,23]. Differences in the characteristics of study populations, occupational categories and study settings might be the reason for these differences.

The association between cervical cancer knowledge and stigma was one of the most important findings of this study. About 80% of women had heard of cervical cancer, but fewer than one third were aware of its signs and symptoms or risk factors. Only 1.6% of participants had heard of HPV in this study. This indicates a considerable gap between general awareness and detailed knowledge. Similar gaps have been documented in Nepal [6,24–27]. A possible explanation for the gap between general awareness and detailed knowledge is the way women receive information about cervical cancer. Many women may have heard about cervical cancer through relatives, friends, or community members, and Female Community Health Volunteers (FCHVs) may also provide some basic information. As a result, women may be generally aware of cervical cancer but may not have a clear understanding of its causes, risk factors, symptoms, prevention, and screening. Since information is not always provided through structured and consistent awareness sessions, some women may remember only the basic message while forgetting the details over time.

Although the present study did not directly examine screening uptake, the observed association between lower stigma and greater cervical cancer-related knowledge suggests that stigma may be relevant to screening-related behaviors [13]. Screening uptake in Nepalese women is influenced by multiple factors beyond stigma, including knowledge, embarrassment, family support, cost, accessibility, privacy, and health-service factors [6,13,24,25]. Therefore, efforts to reduce cervical cancer stigma should be integrated with broader strategies to improve access to and acceptability of cervical cancer screening services.

A key finding of this study was that greater cervical cancer knowledge was associated with lower cervical cancer stigma. Women who knew about cervical cancer, its signs and symptoms, prevention, and treatment reported lower stigma scores. Better knowledge may reduce fear and misconceptions by helping women understand that cervical cancer can be prevented, detected early, and treated. This may reduce feelings of awkwardness and perceptions of cancer as a severe or hopeless disease, as well as concerns about its financial consequences. Similar findings from Paneru et al. showed that stigma related to awkwardness, severity, and financial discrimination was associated with lower cervical cancer screening uptake in Nepal [13]. Therefore, better knowledge may help reduce stigma across different domains and contribute to lower overall stigma.

The lack of a significant association between risk-factor awareness and cervical cancer stigma is noteworthy. Information about risk factors may sometimes be interpreted in terms of personal responsibility or blame, which is also reflected in the personal responsibility domain of the CASS [10]. Studies have also shown that risk-factor information with clear messages about HPV, prevention, screening, and treatment, while emphasizing that cervical cancer can occur despite appropriate preventive behaviors may improve attitude towards cancer [28].

The findings have several implications for cervical cancer prevention and stigma reduction in Nepal. First, cervical cancer education should extend beyond general awareness to include accurate and understandable information on HPV, symptoms, prevention, screening, and treatment. This is particularly important given the persistent knowledge gaps documented among women in Nepal[25–27]. Second, educational approaches may need to be adapted to women’s social and educational backgrounds. Community-based health education has shown promise in Nepal; for example, a community-based educational program in rural Nepal substantially increased women’s willingness to participate in cervical cancer screening following an educational meeting [29]. Third, stigma should be considered alongside knowledge and access when designing screening programs. Qualitative evidence from Nepal has shown that stigma, social norms, lack of awareness, and concerns about screening can influence women’s decisions about screening [6]. However, intervention studies are needed to determine whether improving cervical cancer knowledge actually reduces stigma and whether reductions in stigma subsequently increase screening uptake.

This study has several strengths. First, it used the Nepali version of the CASS, which has demonstrated satisfactory reliability and validity among Nepali women [23]. The scale captures multiple dimensions of cancer stigma rather than relying on a single measure. Second, the study examined a broad range of sociodemographic, reproductive, and cervical cancer-related knowledge characteristics. Third, the use of generalized estimating equations accounted for clustering of participants within wards, which was appropriate given the cluster-based design of the parent study.

However, this study has several limitations. First, the cross-sectional design prevents determination of temporality and causality. Second, knowledge and other characteristics were self-reported and may therefore be subject to recall and social desirability bias. Third, the study was conducted in a single municipality, limiting generalizability to other urban populations in Nepal. Fourth, participants were restricted to women aged 30–60 years who met specific eligibility criteria, so the findings should not be generalized to all women. Finally, residual confounding by factors such as health literacy, previous exposure to cancer among family members, access to health information and healthcare utilization cannot be excluded.

## Conclusion

This study identified important sociodemographic and cervical cancer-related knowledge factors associated with cervical cancer stigma among married women living in an urban municipality of Nepal. Lower stigma was associated with formal education and greater awareness of cervical cancer, including its signs and symptoms, prevention, treatment and screening. Higher stigma was observed among Janjati and other ethnic groups and among homemakers and self-employed women compared with women in salaried employment. These findings highlight the importance of addressing social and informational factors when developing strategies to reduce cervical cancer stigma. Culturally appropriate and accessible information on cervical cancer and screening may be particularly important for groups with lower levels of education or limited access to health information. Longitudinal and intervention studies are needed to determine whether improving cervical cancer knowledge can reduce stigma and whether reductions in stigma contribute to greater uptake of cervical cancer screening.

## Data Availability

All data produced in the present work are contained in the manuscript

## Declarations

## Ethics approval and consent to participate

Ethical approval was obtained from the Institutional Review Committee of Kathmandu University School of Medical Sciences (IRC reference number: 42/22). Written informed consent was obtained from literate participants. For participants unable to read or write, verbal informed consent was obtained and documented through a witness selected by the participant, who signed the consent form. Confidentiality and voluntary participation were maintained throughout the study, and participants could withdraw at any time without any consequences. All methods were carried out in accordance with the Declaration of Helsinki.

## Consent for publication

Not applicable.

## Availability of data and materials

All data generated or analysed during this study are included in this published article and its supplementary information files.

## Authors’ contributions

Priyanka Timsina: Conceptualization, data curation, formal analysis, investigation, methodology, project administration, resources, software, supervision, validation, visualization, writing of the original draft and review and editing of the manuscript.

Bandana Paneru: Conceptualization, formal analysis, methodology, software, supervision, validation and review and editing of the manuscript.

Manjari Shrestha: Data curation, investigation, project administration, resources and writing of the original draft.

Asika Ghemosu: Data curation, investigation, project administration, resources and writing of the original draft.

Bihungum Bista: Formal analysis, methodology, software, supervision, validation, and review and editing of the manuscript.

Archana Shrestha: Conceptualization, data curation, formal analysis, investigation, methodology, project administration, resources, software, supervision, validation, visualization, writing of the original draft and review and editing of the manuscript. All authors read and approved the final manuscript.

## Acknowledgements

The authors thank the Department of Public Health and Community Programs, Kathmandu University School of Medical Sciences and the Budanilkantha Municipality for constant support and guidance throughout the study.

## Supporting Information

S1 STROBE Checklist

S2 Ethics Approval Letter.

S3 Dataset

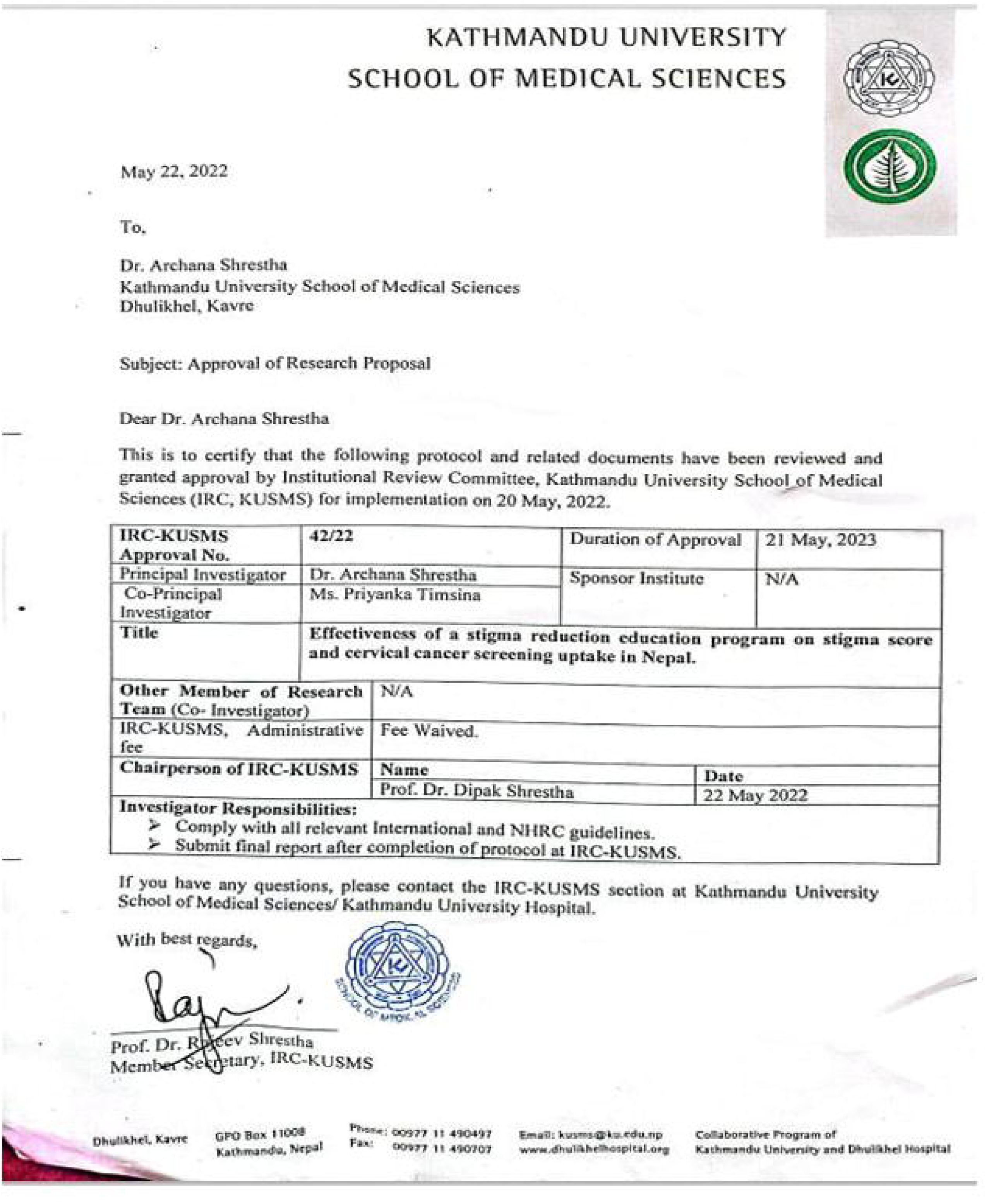

